# Modelling *gambiense* human African trypanosomiasis infection in villages of the Democratic Republic of Congo using Kolmogorov forward equations

**DOI:** 10.1101/2021.05.24.21257532

**Authors:** Christopher N. Davis, Matt J. Keeling, Kat S. Rock

**Affiliations:** Mathematics Institute, University of Warwick, Coventry, CV4 7AL, UK; Zeeman Institute (SBIDER), University of Warwick, Coventry, CV4 7AL, UK; School of Life Sciences, University of Warwick, Coventry, CV4 7AL, UK

**Keywords:** Mathematical Model, Kolmogorov Forward Equations, African Trypanosomiasis, African Sleeping Sickness

## Abstract

Stochastic methods for modelling disease dynamics enables the direct computation of the probability of elimination of transmission (EOT). For the low-prevalence disease of human African trypanosomiasis (gHAT), we develop a new mechanistic model for gHAT infection that determines the full probability distribution of the gHAT infection using Kolmogorov forward equations. The methodology allows the analytical investigation of the probabilities of gHAT elimination in the spatially-connected villages of the Kwamouth and Mosango health zones of the Democratic Republic of Congo, and captures the uncertainty using exact methods. We predict that, if current active and passive screening continue at current levels, local elimination of infection will occur in 2029 for Mosango and after 2040 in Kwamouth, respectively. Our method provides a more realistic approach to scaling the probability of elimination of infection between single villages and much larger regions, and provides results comparable to established models without the requirement of detailed infection structure. The novel flexibility allows the interventions in the model to be implemented specific to each village, and this introduces the framework to consider the possible future strategies of test-and-treat or direct treatment of individuals living in villages where cases have been found, using a new drug.

## Background

In mathematical epidemiology, a growing number of models employ the use of stochastic events to describe infection dynamics. These stochastic methodologies, such as the Gillespie or tau-leaping algorithm, typically use a large number of event-driven stochastic simulations to estimate a distribution of possible behaviours for an epidemic [1]. A central benefit of stochastic models is that the random nature of each realisation means a larger range of outcomes is captured than simply the equilibrium dynamics predicted by a deterministic model [2]. These stochastic methods are also integer-based and so the exact number of people infected at a given time is monitored, including when infections reach zero; a deterministic model will never reach zero infections and so a threshold needs to be applied to reach the elimination [3–5]. Thus, a model in a stochastic framework, will have different expected behaviour and be better suited than a deterministic variant when close the the elimination threshold, either due to low infection numbers or small populations [6].

However, event-driven stochastic methods require a large number of realisations of the epidemic process to be generated to be confident that the full distribution of events has been captured; even then this is still only an approximation of the true probability distribution of the potential trajectories for the infection dynamics in time. This is particularly important if there are any rare events of the system — something that requires more realisations to determine the true frequency at which they occur [6]. Alternative to these event-driven stochastic simulations, the Kolmogorov forward equations provide a method incorporating the stochastic behaviour in a set of ordinary differential equations (ODEs), which fully determine the probability distribution of the epidemic over all possible infection states for the population [6]. This approach is simple to formulate, as many systems are linear in terms of the probability of being in each infection state, and so can be written in a matrix formulation; other methods exist for when this is not the case [7]. The system is thus easy to solve using standard methods and provides a complete description of the dynamics. It is also much faster to solve than repeatedly generating realisations of the stochastic process. The solutions are also numerically exact and an array of derived quantities can be directly calculated.

Kolmogorov forward equations have been utilised in several epidemiological contexts [8–10] and discussed as a powerful tool, but are not widely used to constraints on computer memory [11]. Every possible infection state must be explicitly tracked and so if a population is large (and particularly if each individual can be in any of a large number of infection states), the number of these states quickly becomes prohibitive to the method, as the number of required computations becomes infeasible. Simulation methods take a lot of computation time; Kolmogorov forward equation methods take a lot of memory storage capacity [12]. Therefore, a Kolmogorov forward equation approach to modelling infections dynamics would be most applicable for a disease with a relatively small number of cases that has the potential to achieve elimination [13]. One such disease is *gambiense* human African trypanosomiasis (gHAT).

Infection with gHAT is a caused by a parasite *Trypanosoma brucei gambiense* and is transmitted by tsetse across Central and West Africa, with the majority of infection occurring in the Democratic Republic of Congo (DRC). This infection has been traditionally controlled with active and passive screening, with subsequent treatment of infection individuals and has been targeted for elimination of transmission (EOT) by 2030 by the World Health Organization (WHO). There have been a substantial number of recent modelling studies that use compartmental models for gHAT, with these studies typically using deterministic systems of ordinary differential equations (ODEs) [14–18], with more recent studies also considering stochastic event-driven approaches [19–22].

Here, we have developed new model for gHAT infection in spatial connectivity of villages within health zones of DRC. We use a lower-dimensional state space for infection than considered in the majority of the literature, which hence allows for Kolmogorov forward equations to be used to define the dynamics. This formulation fully captures the possible behaviour of the infection in a stochastic framework, while exhibiting the advantages of a providing the full and exact distribution of the infection states. We investigate the probabilities of gHAT persistence or extinction and compute expected times until elimination of infection, which, despite the lower-dimensional infection structure, provides comparable results to more commonly used methods.

Furthermore, the Kolmogorov forward equation model provides the necessary formulation to explore the interactions between a large number of villages. Village-specific simulations can mimic the real-world interventions observed at the level at which they occur, with the results then realistically scaled up to larger regions, where elimination of infection can be considered at more meaningful geographic scales [23]. In the context of gHAT, the method provides a link between individual village [19] and health zone [18] modelling, while including the stochastic properties required to directly simulate elimination of infection, we can assess potential strategies and progress towards achieving elimination goals.

## Methods

### Kolmogorov forward equations

We construct our Kolmogorov forward equation model by adapting the structure of the suite of gHAT models first presented in Rock *et al*. [14] and updated in Crump *et al*. [17]. Unlike these previous models, the model presented in this manuscript contains just two infection states for a person (susceptible and infected) and two types of people (low- and high-risk of exposure to tsetse bites), and does not explicitly model the number of infected tsetse, the biological vector of the disease.

We derive the new model equations by replacing the tsetse dynamics of previous models with the quasi-equilibrium solution, in order to limit the number of model compartments. For the case gHAT, this assumption is justified by the short life-expectancy of the vector (tsetse) [24] and the long timescales of the infection in humans [25]. The number of infection compartments for humans are also reduced from five (susceptible, exposed, infected Stage 1, infected Stage 2, and hospitalised) to two, whereby exposed, infected Stage 1 and infected Stage 2 are now included as a single infected state, *I*_*H*_, and the former susceptible and hospitalised compartments are now given as all susceptible *S*_*H*_ (Figure 1). The total human population size is constant and denoted as *N*_*H*_, with a small natural mortality rate of people, *µ*_*H*_, replaced by new susceptible individuals. We retain a risk structure whereby a small minority of the population is high-risk, with a higher exposure to biting tsetse and failure to attend active screening.

**Figure 1:**
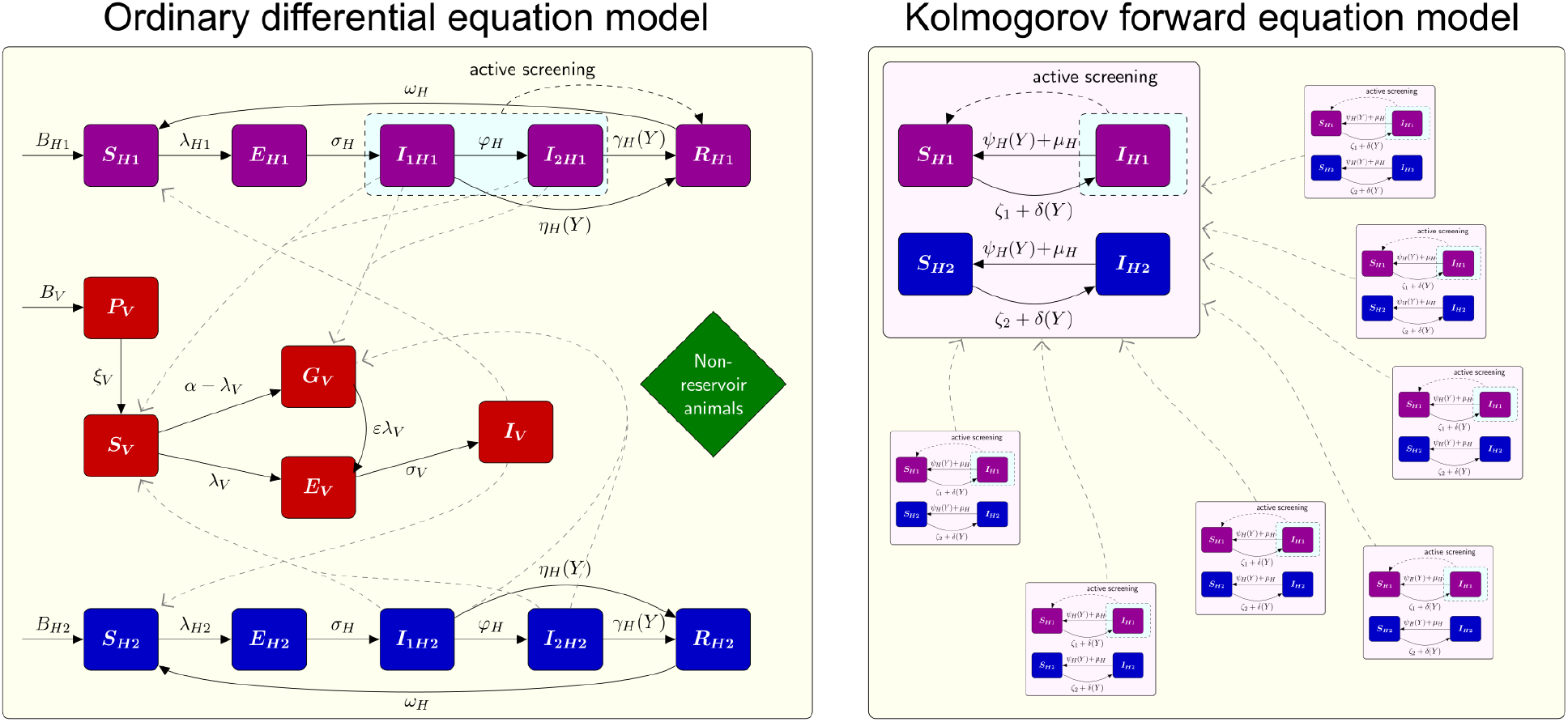
Model diagrams for the two gHAT models. The left panel shows the full higher-dimensional ODE model from Crump *et al*. [17] that includes tsetse dynamics. The right panel shows the lower-dimensional spatially-connected Kolmogorov forward equation model presented in this manuscript. In both panels the solid black lines shows the rate of movement between compartments and dashed black lines shows the the result of active screening and treatment of infected individuals. The dashed grey paths show interaction between humans and tsetse or between all villages, respectively in each panel.

This leaves four possible infection states for any person, and therefore the lower-dimensional model structure, but for an ODE framework is given by Equations 1 and 2.

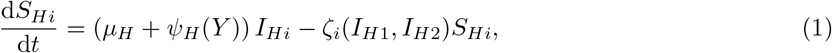

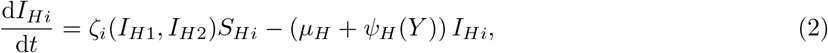

for *i* = 1, 2, for the low- and high-risk group respectively. The natural human mortality rate, *µ*_*H*_, determines the total human birth rate, *µ*_*H*_*N*_*H*_, such that a constant population size is maintained. The recovery rate, *ψ*_*H*_ (*Y*), is dependent on time due to increased detection rates in at later times and is derived from a combination of parameters from Crump *et al*. [17], detailed in the supplementary material. The force of infection is given by *ζ*_*i*_(*I*_*H*1_, *I*_*H*2_), which is a function of the risk class and the number of people infected in each risk class, further depending on the quasi-equilibrium solution for the tsetse (Figure 1). See supplementary information a full explanation of the derivation for these model parameters.

To translate this re-formulated model (Equations 1 and 2) into the Kolmogorov forward equations, we first consider infection state of the population. Since we assume constant population sizes *N*_*H*1_ and *N*_*H*2_ in each risk group and *S*_*Hi*_ + *I*_*Hi*_ = *N*_*Hi*_ for *i* = 1, 2, the infection state is fully determined by the number of infected low- and high-risk people *I*_*H*1_ and *I*_*H*2_. Thus, we define the probability of being in a given state at time *t* as 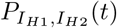. From this population infection state, there are four possible transitions to different states: a low-risk human can recover or die, a high-risk human can recover or die, a low-risk human can get infected, or a high-risk human can get infected (since we assume a constant population size, death is followed by immediate replacement in the susceptible class and so we do not consider this separately to recovery). Therefore, the Kolmogorov forward equations are given by:

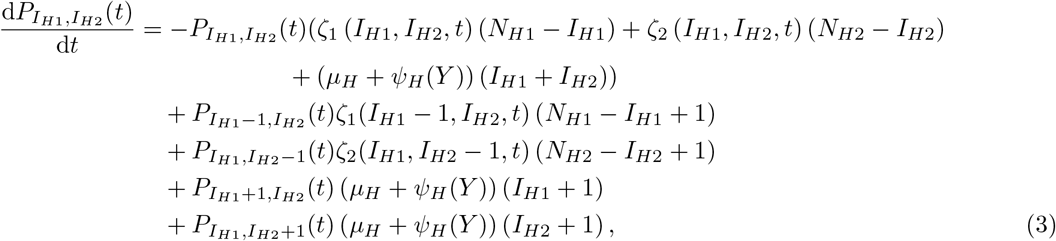

where *I*_*H*1_ = 0, …, *M*_*H*1_ and *I*_*H*2_ = 0, …, *M*_*H*2_. The values *M*_*H*1_ and *M*_*H*2_ are the maximum number of people possible to be infected in each risk group.

In theory, the *M*_*H*1_ = *N*_*H*1_ and *M*_*H*2_ = *N*_*H*2_, however because in practice gHAT is a low-prevalence infection [26], we can reduce the state space of the model and impose a lower maximum threshold, while retaining high model accuracy. We ensure that the probability of exceeding the threshold is very small (less than 1 × 10^−8^), with the values of *M*_*Hi*_ dependent on *N*_*Hi*_, and explicitly given in the supplementary material.

Thus, the Kolmogorov forward equations (Equation 3) comprise of a system of (*M*_*H*1_ + 1)(*M*_*H*2_ + 1) ODEs (since the low-risk infected population can take any value between 0 and *N*_*H*1_ and similarly for the high-risk). The Kolmogorov forward equations are a linear system, and hence we simplify the notation by writing the equations in matrix form. By defining the probability vector:

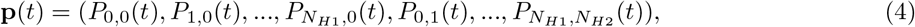

we obtain the Kolmogorov forward equations in matrix form as:

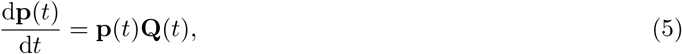

where **Q**(*t*) is the rate matrix of all transition rates at time *t*. We subsequently give the time *t* in both the year *Y* and number of days into that year *d* and hence, **p**(*t*) = **p**(*Y, y*). However, we note that **Q**(*t*) = **Q**(*Y*) because we assume, as per Crump *et al*. [17], that the change in the rate matrix is due to an increase in the rate of passive detection of infection due to improvements in the passive surveillance system, which occurs annually.

Since the equations of Crump *et al*. [17] consider large populations of roughly 100,000 people, rather than much smaller villages, we additionally include a rate of importation of infection into a village, due to movement of people between villages, for which we use a value derived in Davis *et al*. [19] and denote by *δ*(*Y*). The event of an external importation increases the number of infected people in either risk group by one and adds terms to Equation 3 representing a change of state from *I*_*H*1_ low-risk infected and *I*_*H*2_ high-risk by increasing from *I*_*H*1_ − 1 to *I*_*H*1_, increasing from *I*_*H*2_ − 1 to *I*_*H*2_, and increasing from *I*_*H*1_ and *I*_*H*2_ to *I*_*H*1_ + 1 or *I*_*H*2_ + 1 respectively:

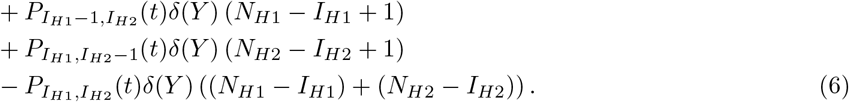

In matrix notation, we include these new terms in matrix **Q**_*E*_(*Y*), whereby the rate matrix is given by **Q**(*Y*) = (**Q**_*V*_(*Y*) + **Q**_*E*_(*Y*)), for the village and external terms respectively.

Additionally, active screening, the process whereby a large number people in a village are targeted to be screened for the disease and then treated if infected, is modelled as a the multiplication of the probability vector **p**(Y,y) by a lower-triangular transition matrix **A**(*Y*). In line with previous modelling studies, we assume that only the low-risk class are affected by this discrete-time event that occurs annually at the beginning of each year, whereas the high-risk class do not attend active screening events. The active screening matrix **A**(*Y*) is calculated, for a given screening coverage (which can change each year), by the use of a hypergeometric distribution to determine the number of infected people screened, followed by a binomial distribution to find the number of infections detected due to the imperfect sensitivity of the test (full details are in the supplementary material).

We assume that active screening began in 1998 [27], and the system was previously at endemic equilibrium given there had been no screening for several years previously. Therefore, we derive the full distribution of infection states for a population in day *d* of year *Y* as

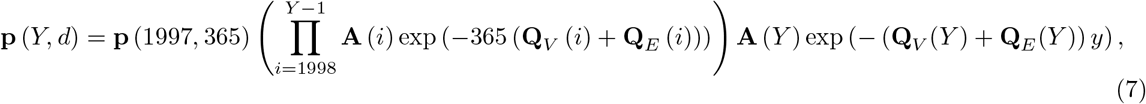

for *Y ≥*1998 and 0 ≤*d* ≤365. Active screening in year *Y* is modelled as occurring at the start of the year (*d* = 0), and hence the probability state just before active screening in year *Y* is given by **p** (*Y* −1, 365). We calculate **p** (1997, 365) by finding quasi-stationary equilibrium, finding the eigenvector corresponding to the largest eigenvalue of the rate matrix, (**Q**_*V*_ (1997) + **Q**_*E*_ (1997)).

### Parameter values

The parameters are taken from Crump *et al*. [17] and transformed into the new parameters of the Kolmogorov forward equations (see supplementary material). The values of the original parameters are either fixed where well-defined in the literature, and specific to DRC, or as the median estimate of the posterior from fitting to screening and incidence data in the DRC health zone of Kwamouth, using a Metropolis–Hastings MCMC algorithm. The data which span 2000–2016 came from the WHO HAT Atlas [26].

The health zone of Kwamouth is used to parameterise the model since it is a comparatively high-incidence region of the DRC, the country with most cases [28] and pre-2018 (the study period with available data) had no large-scale vector control [29], which would affect the infection dynamics. We additionally present similar results for the low-incidence health zone of Mosango when comparing health zone (Figure 5) and in the supplementary material. We note that the methodology presented here could alternatively be applied to any other health zone or region.

## Results

### Infection dynamics of a single village

Solving the Kolmogorov forward equations, using the matrix exponential in the form presented in Equation 7, we can obtain projections of how the infection dynamics will change in time in full probabilistic form. As an illustrative example, we calculate the distribution of infection up to the year 2030 for a village of 1,000 people in the Kwamouth health zone, assuming that within the village there is an active screening coverage of 50% every year (Figure 2a). We are assuming a small rate of infectious importations from movement of people that decreases with time, *δ*(*Y*) = (3.4 ×10^−6^) exp(− 0.1071(*Y* − 2000)) days^−1^, and that the village starts at endemic equilibrium conditions in 1998, calculated as the steady state of the Kolmogorov forward equations.

**Figure 2:**
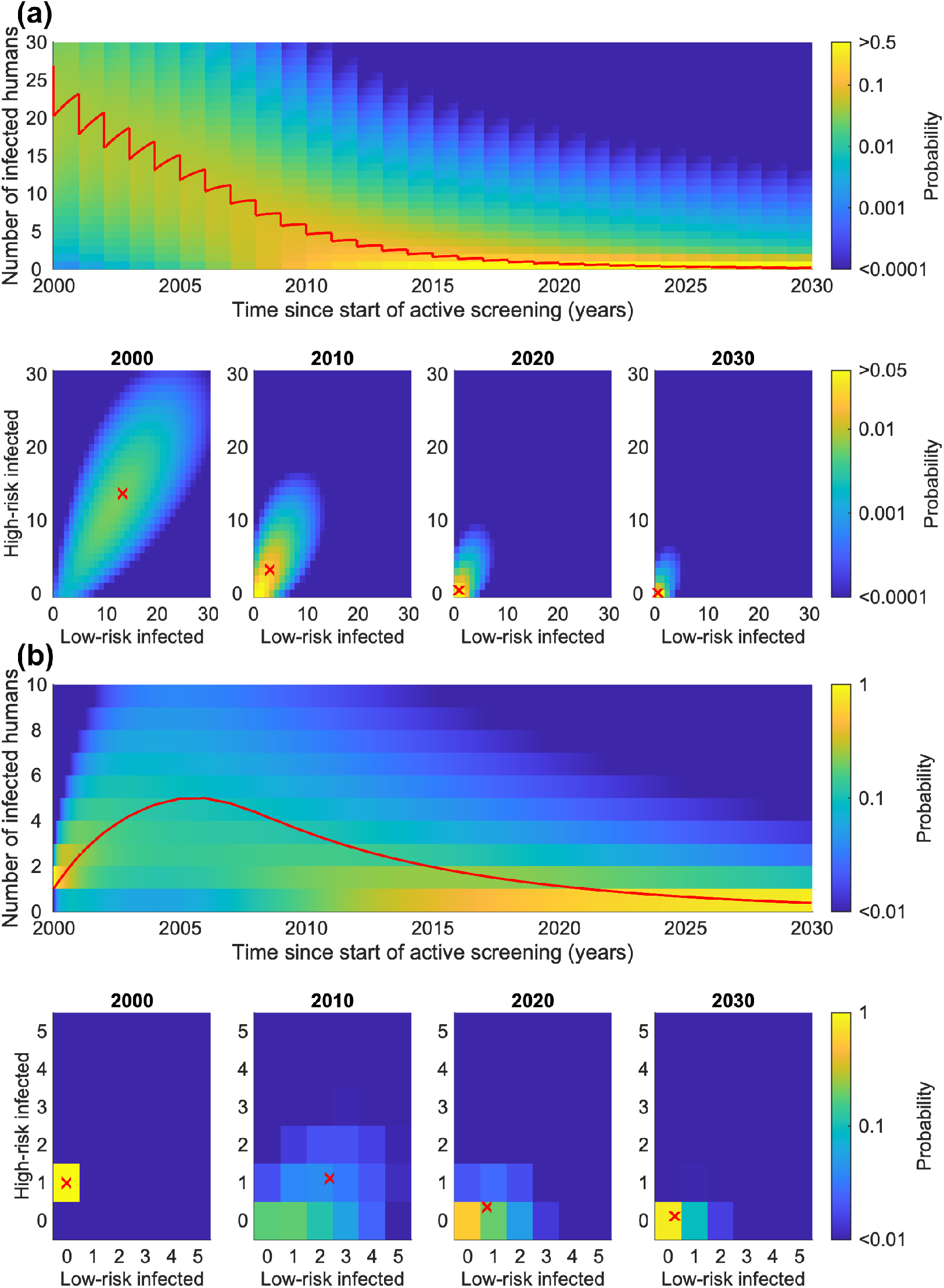
Illustrative example of the distribution of infection in a single example village of population size *N*_*H*_ = 1, 000 for two scenarios. (a) A gHAT-endemic village with 50% annual active screening coverage and a small rate of infectious importations. (b) A village with local elimination of infection pre-2000 followed by the introduction of one high-risk infected person in 2000, including a small rate of infectious importations in the future, but no active screening. For each scenario, the top panel shows the distribution of the total number of infected people in time and the lower panels show the distribution between the risk groups at selected time points. The red line in the top panel shows the mean expected dynamics and the red crosses in the bottom panels shows the expected numbers in each risk class at that time.

The results show that the introduction of active screening drives down the expected number of infected people from the steady-state distribution in 1998. By 2030, there is a probability of 0.90 that there are no infected people in this example village. The steady-state is concentrated on negligible infection levels assuming the active screening is maintained. The full probability distribution does, however, show that for this individual village there is still some probability the infections will not fall as quickly, or indeed remain constant or increase.

These results are dependent on the import rate between villages. Without the importation rate, there is an absorbing disease-free population state dynamics, which the population would eventually move towards (a steady-state of no infection). This is because without the importation rate, if infection levels reach zero there is no person to re-introduce infection the population (albeit through tsetse in practice).

Conversely to an endemic village, we consider a hypothetical village in Kwamouth that was infection-free in 1998 and therefore not targeted for active screening (Figure 2b). We predict some level of resurgence on average in the village if a single high-risk person of the village became infected through travelling to another village. There is a high probability of onward transmission in the village; the expected number of infected people increases initially, before decreasing again. However, infection is unlikely to be maintained in the long term, even without additional controls — there is a probability of 0.83 of a return to no infection by 2030.

### Infection dynamics across multiple villages

The infection dynamics in individual villages informs us about the probability of local elimination of infection for an average village, but at a larger scale, such as health zone or country level, elimination of infection will depend on more than the probability of elimination in individual villages. While the distribution of gHAT infection is heterogeneous across the DRC [30], highly-clustered incidence means that local movement of people will affect the probability of elimination in neighbouring villages. The rate of infectious importations in a village will be dependent on the total infection level in the area.

Therefore, to consider the dynamics across multiple villages in a region, we modify our rate of infectious importations to remove the exponential decrease in time matched to the trend in global infections and replace this with a term proportional to the total number of expected infections in model predictions across all villages of the local study region — in this case the health zone of Kwamouth — such that *δ*(1998) = (3.4 ×10^−6^) days^−1^. The supplementary material provides a complete description of how the rate of infectious importations is formulated.

We consider the expected number of infections and the probability of elimination of infection for four groups of 10,000 people, comprised of groups of villages of different sizes (*N*_*H*_ = 10, 000, 1, 000, 100 and 10) (Figure 3) to understand the impact of the metapopulation structure [23]. The smaller village population sizes within total populated area have fewer expected gHAT infections and a higher probability of elimination of infection. The reduced number of interactions of mixing in smaller villages also results in a lower steady-state from before the active screening begins. There is a much lower probability of elimination of infection when there are fewer larger villages. For 1,000 villages with a population size *N*_*H*_ = 10, the probability of elimination of infection in 2030 is > 0.99, while for just one village of *N*_*H*_ = 10, 000, there is a smaller probability of 0.77 for elimination of infection by 2030. This is in agreement with results of metapopulation studies, where more stochastic fade-outs of infection occur in the smaller populations, leading to a greater probability of elimination of infection across larger areas when sub-divided into more populations [31]. The example here highlights the benefit of modelling the full stochastic dynamics, where the small population sizes determine the frequency of extinction events; the results from an ODE model with constant population size would not vary with the size of the population.

**Figure 3:**
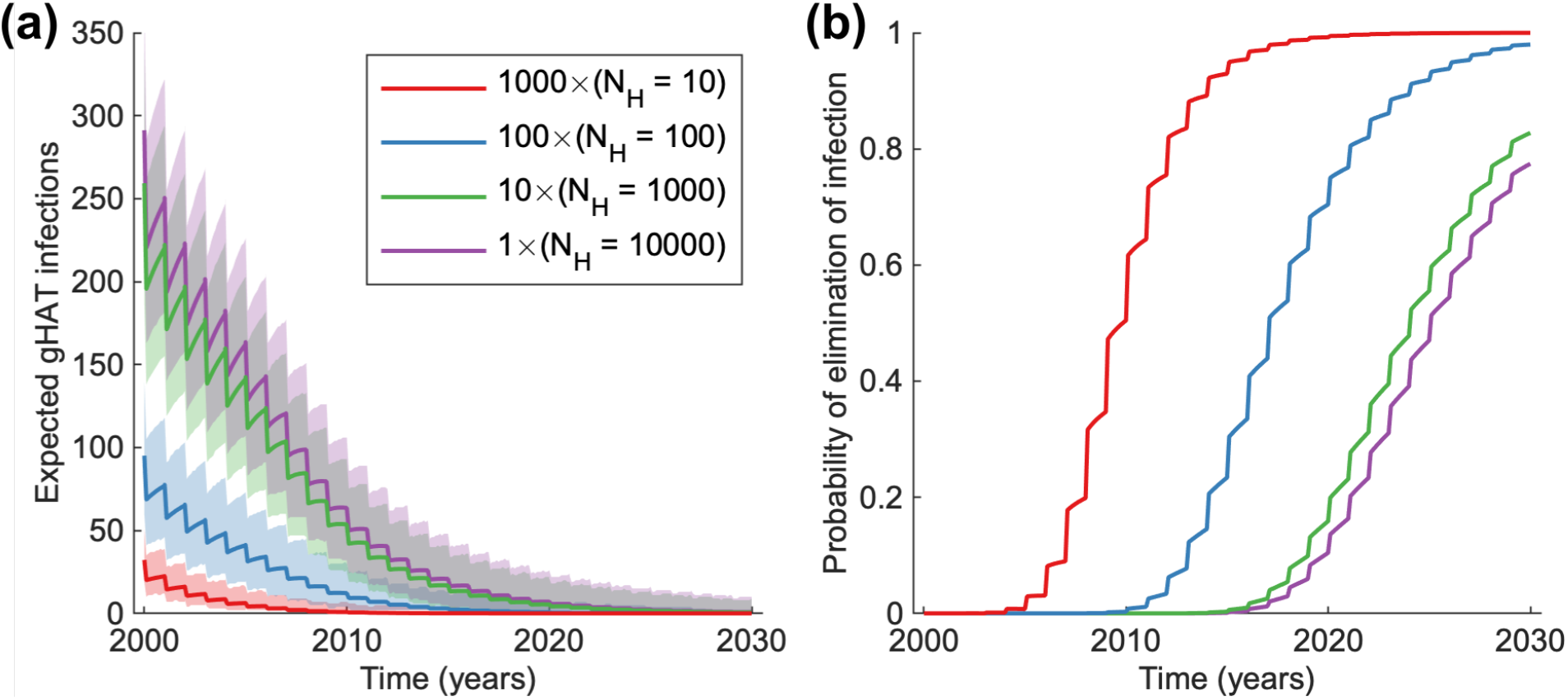
Infection dynamics in a total population of 10,000, partitioned into groups of villages of different sizes *N*_*H*_ = 10, 000, 1, 000, 100 and 10. The annual active screening coverage is 50% and projections are started for the equilibrium distribution in 1998. (a) The expected number of infections. The shaded region shows the 95% prediction intervals around the solid line for the expected value. (b) The probability of local elimination of infection.

### Infection dynamics across a health zone

While we have considered theoretical villages to explore the behaviour of the Kolmogorov forward equation model, we now use data from the WHO HAT Atlas [28] to obtain a list of real villages to apply our model to. Since our model uses parameters matched to the data from the health zone of Kwamouth, we extract the population sizes of each village in Kwamouth (see supplementary material for details) along with the past active screening coverage. A plausible screening pattern is obtained taking the mean active screening coverage across all screenings and the probability that any particular village listed in the WHO HAT Atlas is screened in a given year. For Kwamouth, these values are a 68.6% coverage occurring at a probability of 0.23 each year using active screening data from 2000–2018 (see supplementary material for details). This active screening scheme is incorporated into the model by a new parameter for the probability of an active screening event in a village. Thus, we model active screening as the linear combination of the probability of no screening multiplied by the current distribution and the probability of a screening multiplied by the distribution after an active screening of given coverage.

We additionally adapt the value of the rate of importations of infection at steady state (previously taken from Davis *et al*. [19]). We calculate the value that is now specific to the health zone of Kwamouth as *δ*(*Y*) = 2.86 × 10^−6^ days^−1^, which is determined by matching the steady state of the whole health zone to the steady state of the system of ODEs in the original model for Kwamouth.

Applying the full Kolmogorov forward equation model to the health zone of Kwamouth, we obtain a full probability distribution in time for each village. We present the probability distribution of infection across the risk groups for a selection of villages of different sizes (*N*_*H*_ = 100, 971, 5, 628 and 20, 697) and for the whole health zone at key time points (Figure 4). Similar results for the lower-endemicity health zone of Mosango are presented in the supplementary material.

**Figure 4:**
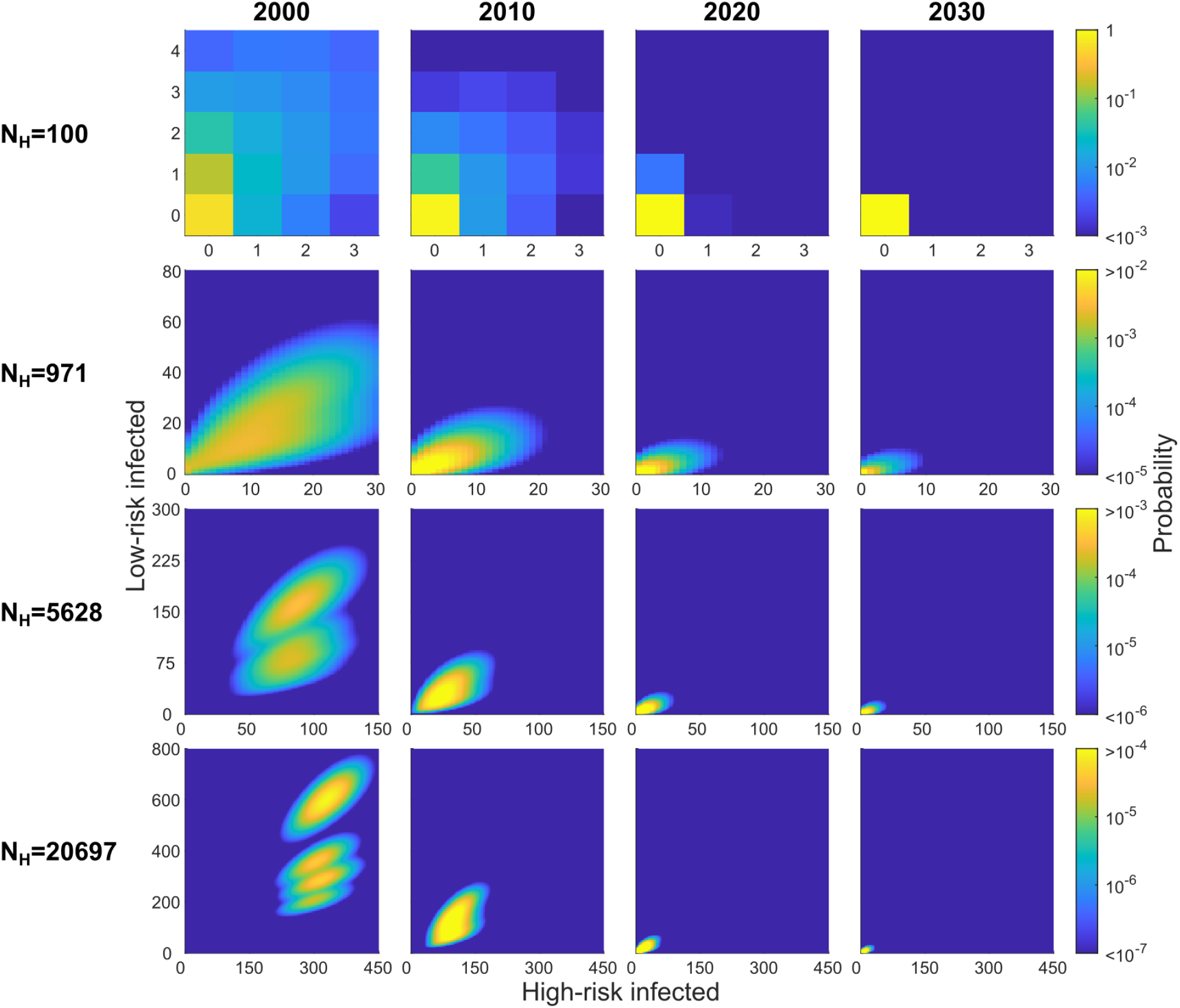
The risk distribution of infection in selected villages of Kwamouth at different time points (the years 2000, 2010, 2020 and 2030); the first possible active screening of each village was in 1998. There are 418 villages with populations ranging between 3 and 20,697 of which we present the probability distribution of infected people in four villages (*N*_*H*_ = 100, 971, 5, 628 and 20, 697). On each subplot the *x*-axis represents the number of high-risk people infected, and the *y*-axis, the number of low-risk people infected. The risk distribution of infection for the alternative health zone of Mosango are presented in the supplementary material.

The expected number of infections in all villages decreases in time, such that by 2030 most of the probability is centred around no infection. For the smallest villages, there is a large probability of local gHAT elimination by 2030 (> 0.99 for the village of size *N*_*H*_ < 100) as there are initially few or no cases, which are then identified by active screening, or passive screening and treatment or death. However, for the largest villages, such as the one with a population of *N*_*H*_ = 20, 697, there is a high probability of continued infection with a probability of just 0.26 that elimination of infection will be met in that village by 2030. Hence, we will frequently see local elimination events, with global persistence across the health zone [31].

Active screening is shown to reduce the infection. This is visually explicit in the column for 2000, where there are separate high probability clouds for whether an active screening has occurred and hence the number of infected people in the low-risk group identified and treated (Figure 4). At the beginning of 2000, active screenings may have occurred in both 1998 and 1999 and so in the bottom left panel of Figure 4 (village of population size *N*_*H*_ = 20, 697), is is clear that there are four possible outcomes highlighted as separate regions of the probability distribution: an active screening event occurred in both 1998 and 1999, just 1998, just 1999, or neither 1998 or 1999. This behaviour is less obvious in some smaller villages as the high probability regions overlap, yet is still present. We note that since active screening is identifying only low-risk individuals, infection is being pushed down but proportionally most of the reduction is in the low-risk group.

The decline in expected infection in all of individual villages is also evident total infection of the health zone (Figure 5). This is calculated as the sum of the expected infection in each village. Note that here we do not present the number of cases, but the underlying infections — case numbers would be substantially lower as there are typically high levels of under-reporting [32]. By 2030, the expected number of infected people have greatly decreased, yet persist in low number. This is mirrored in the probability of elimination of infection, calculated as the product of achieving zero infections in all villages of the health zone, which is less than 10^−4^ by 2030 if this active screening coverage remains constant and only identifies low-risk infected individuals, with the mean expected year of elimination after 2040. Using parameter values in the model matched to WHO HAT Atlas data for the low-incidence health zone of Mosango, we observe an earlier mean expected year of elimination of infection in 2029.

**Figure 5:**
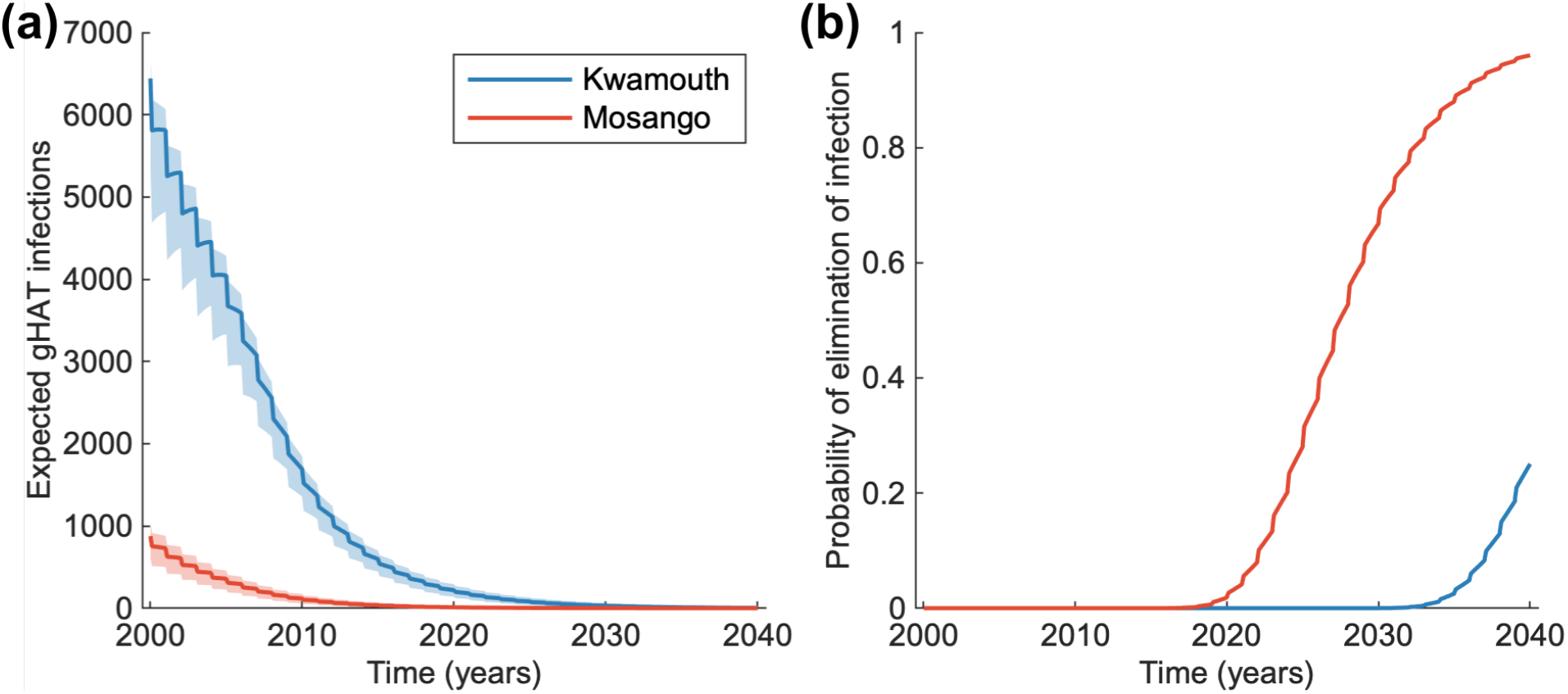
The total infection in the health zones of Kwamouth and Mosango. We assumed Kwamouth had a population of 206,135 people made up 481 distinct villages, while Mosango had a population size of 107,685 of 204 villages. (a) The expected number of infections across all villages in time. The shaded region shows the 95% prediction intervals. (b) The probability of zero infections in time and hence elimination for all villages in each health zone together.

## Discussion

The Kolmogorov forward equation model facilitates a powerful and efficient way to analyse the dynamics of a low-prevalence infections such as gHAT. This method presented here, utilising a lower-dimensional model structure than commonly used models, is fast to compute (with sufficiently small populations) and yet maintains a good correspondence with more complex approaches. The nature of the implementation means that various interesting properties can easily be explored, with exact methods for calculating extinction times and expected dynamics [6]. This approach has also allowed the model to be easily extended to consider the interaction of multiple villages and even to consider the dynamics of persistence at the health zone level by linking the total number of infected individuals to the rate of infectious imports into the villages.

Using this model, we conclude that based upon the strategy of active screening at the mean level, the expected year of elimination of infection is after 2040 for Kwamouth and in 2029 for Mosango. This is in line with deterministic predictions using this parameterisation but at a health zone level; Huang *et al*. [18] predicted that the year of elimination of infection would be after 2040 for Kwamouth and in 2031 for Mosango, using a similar model and the mean coverage of active screening. Likewise, stochastic models at a health zone level, found that without COVID-19 interruptions to gHAT activities, elimination might be expected in 2025 in Mosango (with this slightly earlier prediction may be explained by higher assumed screening from 2017 onwards) [21]. This general agreement between either connected village scale and health zone scale models is reassuring and supports continued use of both model frameworks, however by formulating connected village-scale methods it is possible to consider spatial dynamics in a more nuanced way.

We note that in considering the populations, we have only used villages that are listed in the WHO HAT Atlas and there are known to be additional villages within these health zones that are not listed, since they may not have ever been screened. Hence, the distribution of active screening is not truly as uniformly distributed across the health zones as presented here. As shown in the supplementary material, we also know that screening coverage and frequency is correlated with population size of a settlement, and so adapting our model to account for this would also likely result in more accurate predictions. The proportion of people in each risk class is also constant for all population sizes in the model, whereas we could speculate that the larger populations are more town-like and perhaps have fewer high-risk members. This potential over-estimation of high-risk people in the large populations could explain some of the very high gHAT persistence probabilities seen in results for these populations (Figure 4).

The assumption that movement of individuals is proportional to the expected number of people infected in the health zone, rather than the full distribution, is also a simplification to avoid calculating all possible combinations infection states in each village. This is only true for a large number of villages, but hence a valid assumption at the health zone level.

In addition, we consider a probability of active screening every year, despite the fact that continued active screening is unlikely to be necessary for small villages, where the infection is almost certain to be locally eliminated in latter years. To improve the plausibility of the model, we could add a cessation criterion, similar other studies [20, 33, 34]. This is less straightforward to implement in this probabilistic framework than in the tau-leaping scheme, as we do not consider specific realisations of the model where the infection is either detected or not, but have a full probability distribution of all possible infection states. One potential solution could be to link the probability of being screened to the probability of observing a case in active screening. We do show that the assumption of a single screening event each year, as opposed to continuously throughout the year, shows negligible differences and so adopt this method for simplicity (see supplementary material).

We have no data on the movement of people between villages, and so the rate of importation was estimated by matching to the probability of detecting infection on the first active screening in a village [19] or matching for the health zone to the expected equilibrium state of an ODE model variant [18]. However, using these values as an approximation for the mixing between villages, we achieve a good match to other model variant (see supplementary material), while retaining the efficiency of the Kolmogorov forward equations and the additional benefits of calculating the full probability distribution.

In the future, this modelling approach could be very valuable for assessing not only decisions about continuation or cessation of screening in specific villages (based on village size and previous detections) but also provides a method through which the impact of village-level mass drug administration on health zone transmission dynamics could be assessed. Whilst such as drug is not currently licensed for this type of delivery, a new compound – acoziborole – which is in phase 3 clinical trials as a single-dose cure, is a possible candidate [35]. These types of village-scale strategy decisions would be challenging to analyse through health zone level approaches.

## Conclusions

We have shown that a lower-dimensional model of gHAT that operates at the village level can achieve very similar results to a more biologically realistic version for larger spatial scales, while introducing a method of obtaining numerically exact results for extinction times and expected number of infected individuals. The predictions provided are in line with previous deterministic and stochastic results and the model implementation provides a framework to scale between modelling at a health zone or national level and at the village level. This is an important development, since the data obtained and the actual interventions (active screening) conducted are at village level.

The Kolmogorov forward equation model suggests that with mean coverage of active screening and continuing passive screening, with no additional interventions such as vector control, the infection is almost certain to persist for long periods. This indicates that additional or intensified controls are required to achieve elimination of transmission, such as tsetse control, improved passive detection or targeting of high-risk people in active screening. This model structure expands the range of analytical projections it is possible to generate and demonstrates the results that can be obtained with a Kolmogorov forward equation model.

## Data Availability

Epidemiological data for the study were provided by the WHO in the frame of the Atlas of gHAT which may be viewed at www.who.int/trypanosomiasis_african/country/risk_AFRO/en and may be requested through Jose Ramon Franco (francoj{at}who.int).

## Acknowledgments

The authors thank Programme National de Lutte contre la Trypanosomiase Humaine Africaine (PNLTHA) of DRC and its director Dr Erick Mwamba Miaka for original data collection and WHO for data access (in the framework of the WHO HAT Atlas [28]).

