## Appendix for "Modelling *gambiense* human African trypanosomiasis infection in villages of the Democratic Republic of Congo using Kolmogorov forward equations"

#### Adapting the ODE gHAT model to Kolmogorov forward equations

The *gambiense* human African trypanosomiasis (gHAT) Kolmogorov forward equations model presented in the main manuscript is given by the equations:

$$\begin{aligned}\frac{\mathrm{d}P_{I_{H1},I_{H2}}(t)}{\mathrm{d}t} = &-P_{I_{H1},I_{H2}}(t)(\zeta_1\left(I_{H1},I_{H2},t\right)\left(N_{H1}-I_{H1}\right)+\zeta_2\left(I_{H1},I_{H2},t\right)\left(N_{H2}-I_{H2}\right)\\ &\qquad+\left(\mu_H+\psi_H(Y)\right)\left(I_{H1}+I_{H2}\right))\\ &+P_{I_{H1}-1,I_{H2}}(t)\zeta_1(I_{H1}-1,I_{H2},t)\left(N_{H1}-I_{H1}+1\right)\\ &+P_{I_{H1},I_{H2}-1}(t)\zeta_2(I_{H1},I_{H2}-1,t)\left(N_{H2}-I_{H2}+1\right)\\ &+P_{I_{H1}+1,I_{H2}}(t)\left(\mu_H+\psi_H(Y)\right)\left(I_{H1}+1\right)\\ &+P_{I_{H1},I_{H2}+1}(t)\left(\mu_H+\psi_H(Y)\right)\left(I_{H2}+1\right),\end{aligned} \tag{1}$$

where $P_{I_{H1},I_{H2}}(t)$ is the probability of there being $I_{H1}$ infected people classed as low-risk people and $I_{H2}$ infected people classed as high-risk people. The same model structure could alternatively be expressed by an ordinary differential equation variant:

$$\frac{\mathrm{d}S_{Hi}}{\mathrm{d}t} = \left(\mu_H+\psi_H(Y)\right)I_{Hi}-\zeta_i(I_{H1},I_{H2})S_{Hi}, \tag{2}$$

$$\frac{\mathrm{d}I_{Hi}}{\mathrm{d}t} = \zeta_i(I_{H1},I_{H2})S_{Hi}-\left(\mu_H+\psi_H(Y)\right)I_{Hi}, \tag{3}$$

for $i = 1, 2$. The models consider two types of people (low- and high-risk are given as $i = 1$ and 2 respectively), that can either be susceptible ($S_H$) or infected ($I_H$).

The model design and parameterisation has been adapted from the model described as 'Model 4', which was first presented in Rock et al. [1], and updated in Crump et al. [2]. The model in Crump et al. [2] is a mechanistic compartmental model for gHAT infection that further divides people of each risk class into five compartments defined as: susceptible $S_H$, exposed (but not infectious) $E_H$, Stage 1 infection $I_{1H}$, Stage 2 infection $I_{2H}$ and hospitalised (or recovering at home) $R_H$. The model outputs the number of humans in each compartment and the proportion of the total number of vectors in each infection compartment, where the vectors (tsetse) are: in the pupal stage $P_V$, teneral (unfed) $S_V$, infected and in their extrinsic incubation period (there are three classes to create a gamma distributed period) $E_V$, infectious tsetse $I_V$, and non-teneral (fed) but uninfected tsetse $G_V$. The population size was assumed to be constant and thus $N_{Hi} = S_{Hi}(t) + E_{Hi}(t) + I_{1Hi}(t) + I_{2Hi}(t) + R_{Hi}(t)$, $i = 1, 2$, where $i = 1$ for low-risk individuals, randomly participating in active screening, and $i = 2$ for high-risk individuals, never participating in active screening, each at time $t > 0$. The compartments of the model [1, 2] and the possible transitions between them are shown graphically in Figure S1, where the transmission of infection between humans and tsetse is shown by grey paths. The full ODE model equations are given in Equations 4–16:

$$\frac{\mathrm{d}S_{Hi}}{\mathrm{d}t} = \mu_H N_{Hi} + \omega R_{Hi} - \alpha m_{\text{eff}} f_i \frac{S_{Hi}}{N_{Hi}} I_V - \mu_H S_{Hi} \tag{4}$$

$$\frac{\mathrm{d}E_{Hi}}{\mathrm{d}t} = \alpha m_{\text{eff}} f_i \frac{S_{Hi}}{N_{Hi}} I_V - (\sigma_H + \mu_H) E_{Hi} \tag{5}$$

$$\frac{\mathrm{d}I_{1Hi}}{\mathrm{d}t} = \sigma_H E_{Hi} - (\varphi_H + \eta_H(Y) + \mu_H) I_{1Hi} \tag{6}$$

$$\frac{\mathrm{d}I_{2Hi}}{\mathrm{d}t} = \varphi_H I_{1Hi} - (\gamma_H(Y) + \mu_H) I_{2Hi} \tag{7}$$

$$\frac{\mathrm{d}R_{Hi}}{\mathrm{d}t} = \eta_H(Y) I_{1Hi} + \gamma_H(Y) I_{2Hi} - (\omega_H + \mu_H) R_{Hi} \tag{8}$$

$$\frac{\mathrm{d}P_V}{\mathrm{d}t} = B_V N_V - (\xi_V + \frac{P_V}{K}) P_V \tag{9}$$

$$\frac{\mathrm{d}S_V}{\mathrm{d}t} = \xi_V \mathbb{P}(\text{pupating}) P_V - \alpha S_V - \mu_V S_V \tag{10}$$

$$\frac{\mathrm{d}E_{1V}}{\mathrm{d}t} = \alpha p_V (f_{H1} \frac{I_{1H1} + I_{2H1}}{N_{H1}} + f_{H2} \frac{I_{1H2} + I_{2H2}}{N_{H2}})(S_V + \varepsilon G_V) - (3\sigma_V + \mu_V) E_{1V} \tag{11}$$

$$\frac{\mathrm{d}E_{2V}}{\mathrm{d}t} = 3\sigma_V E_{1V} - (3\sigma_V + \mu_V) E_{2V} \tag{12}$$

$$\frac{\mathrm{d}E_{3V}}{\mathrm{d}t} = 3\sigma_V E_{2V} - (3\sigma_V + \mu_V) E_{3V} \tag{13}$$

$$\frac{\mathrm{d}I_V}{\mathrm{d}t} = 3\sigma_V E_{3V} - \mu_V I_V \tag{14}$$

$$\frac{\mathrm{d}G_V}{\mathrm{d}t} = \alpha(1 - p_V (f_{H1} \frac{I_{1H1} + I_{2H1}}{N_{H1}} + f_{H2} \frac{I_{1H2} + I_{2H2}}{N_{H2}})) S_V \tag{15}$$

$$- \alpha p_V (f_{H1} \frac{I_{1H1} + I_{2H1}}{N_{H1}} + f_{H2} \frac{I_{1H2} + I_{2H2}}{N_{H2}}) \varepsilon G_V - \mu_V G_V, \tag{16}$$

for $i = 1, 2$ (low- and high-risk people).

As described in the main manuscript, the Kolmogorov forward equation model reduces complexity by replacing the explicit tsetse compartments of the model with a quasi-equilibrium solution. This is obtained by solving the tsetse dynamic equations (Equations 9–16) at steady state. This gives the solution:

$$I_V^Q = \frac{27\sigma_V^3 \alpha p_V \left( f_{H1} \frac{(I_{1H1}+I_{2H1})}{N_{H1}} + f_{H2} \frac{(I_{1H2}+I_{2H2})}{N_{H2}} \right) N_H \left(\mu_V + \epsilon\alpha\right)}{\left(\alpha + \mu_V\right) \left( \alpha p_V \left( f_{H1} \frac{(I_{1H1}+I_{2H1})}{N_{H1}} + f_{H2} \frac{(I_{1H2}+I_{2H2})}{N_{H2}} \right) \epsilon + \mu_V \right) \left(3\sigma + \mu_V\right)^3}, \tag{17}$$

which is a function of the number of infected humans. To develop the Kolmogorov forward equations, this term replaces $I_V$ in the reduced set of Equations 4–8.

In the more detailed model, humans can become exposed, $E_H$, upon a bite from an infective tsetse and progress to Stage 1 infection, $I_{H1}$, before either moving to Stage 2 infection $I_{H2}$ followed by the non-infectious class $R_H$, or moving directly to the non-infectious class, if the infection is detected early. Stage 1 detection is at rate $\eta_H(Y) = \eta_H^{\text{post}}(\eta_{H\text{amp}}/(1+ \exp(-d_{\text{steep}}(t - d_{\text{change}}))))$ and Stage 2 is at rate $\gamma_H(Y) = \gamma_H^{\text{post}}(\gamma_{H\text{amp}}/(1 + \exp(-d_{\text{steep}}(t - d_{\text{change}}))))$. The birth rates $B_{Hi}$ are given by $\mu_H N_{Hi}$ for $i = 1, 2$ and $B_V$ is equal to $\mu_V N_H$, where the different numbers of humans and vectors are accounted for in the effective tsetse density $m_{\text{eff}}$.

We remove much of this structure by combining the exposed and infected compartments from each risk class into one compartment (infected) and the susceptible and recovered compartments into one compartment (susceptible). We keep the mean time spent in each compartment (or group of compartments) the same in both models by introducing new parameter $\psi_H$ and new function $\zeta$, which is a combination of previous parameters and the number of infected people. This describes the movement between the new compartments in the reduced model. These are given by:

$$\psi_H(Y) = \frac{1}{1/\sigma_H + \frac{1}{\frac{1}{1/\varphi_H + 1/\gamma_H(Y)} + 1/\eta_H(Y)}} \tag{18}$$

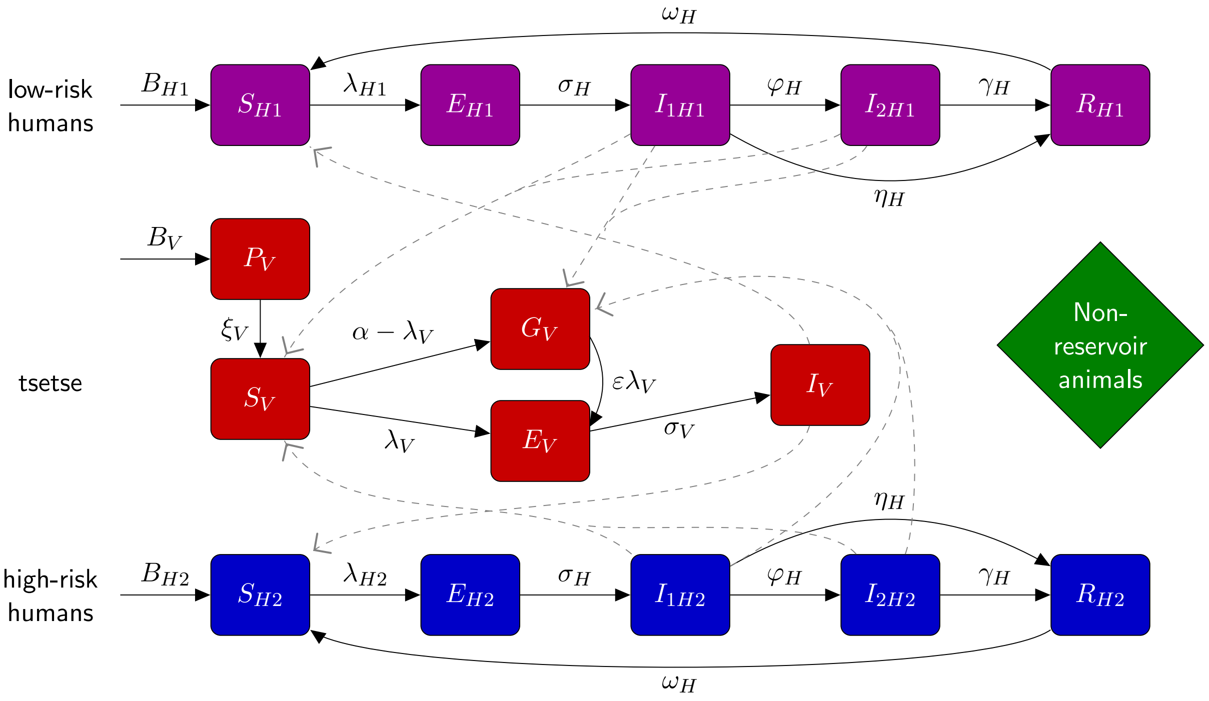


Figure S1: Schematic of the compartmental model for gHAT infection dynamics in humans (low- and high-risk) and tsetse. Adapted from Crump *et al.* [2].

and

$$\zeta_i(I_{H1}, I_{H2}) = \frac{1}{1/\omega_H + \frac{\mu_V + p_V \alpha \epsilon \left( f_{H1} \frac{I_{H1}}{N_{H1}} + f_{H2} \frac{I_{H2}}{N_{H2}} \right)}{\alpha m_{\text{eff}} I_V^Q \left( f_{H1} \frac{I_{H1}}{N_{H1}} + f_{H2} \frac{I_{H2}}{N_{H2}} \right)} \left( \frac{f_{Hi}}{N_{Hi}} \right)} \qquad (19)$$

for $i = 1, 2$. $I_{H1}$ is now defined as the number of low-risk infected people (of either infection stage) and likewise $I_{H2}$ is for high-risk. These definition in Equations 2 and 3 provide the model new structure, while ensuring the the mean time spent in the compartments remains the same as the full model. Values of these original parameters is given in Table S1. These parameter values are taken from Crump *et al.* [2], which were either sourced from literature, where well-defined, or otherwise (in the case of $R_0$, $k_1$, $k_2$, $r$, $u$, Se, $\eta_H^{\text{post}}$, $\eta_{H\text{amp}}$, $\gamma_H^{\text{pre}}$, $\gamma_H^{\text{post}}$, $\eta_{H\text{amp}}$, $d_{\text{change}}$, and $d_{\text{steep}}$) taken as the median of the distribution obtained by model fitting using a Metropolis–Hastings MCMC algorithm that matched the deterministic version of the model to incidence data from the WHO HAT Atlas [3].

#### Reducing the state space

Despite reducing the possible state space in the Kolmogorov forward equations by only including four human model compartments, which is fully determined by just two ($I_{H1}$ and $I_{H2}$) due to the constant population size, the state space is still very large. Therefore, since gHAT is an infection with typically low levels of infection [3], we can reduce the state space by only modelling low levels of infection. It is exceedingly improbable that near the full population would become infected with gHAT.

We introduce $M_{H1}$ and $M_{H2}$ as the maximum number of people possible to be infected in each risk group. In theory, $M_{H1} = N_{H1}$ and $M_{H2} = N_{H2}$; however, we set these values such that there is a probability of less than $1 \times 10^{-8}$ that the infection levels will exceed these numbers at the initial condition. When the number of infected people is equal to these imposed maxima, there is then zero probability of an additional person becoming infected in the model. To ensure the probability of such an event remains small, the values of $M_{H1}$ and $M_{H2}$ dependent on $N_H$ (given in Figure S2). For the largest village in Kwamouth, with a population size of 20,697, it is only necessary to model up to $M_{H1} = 1,084$ and $M_{H2} = 594$ to ensure that the probability of infected people exceeding these numbers is less than $1 \times 10^{-8}$.

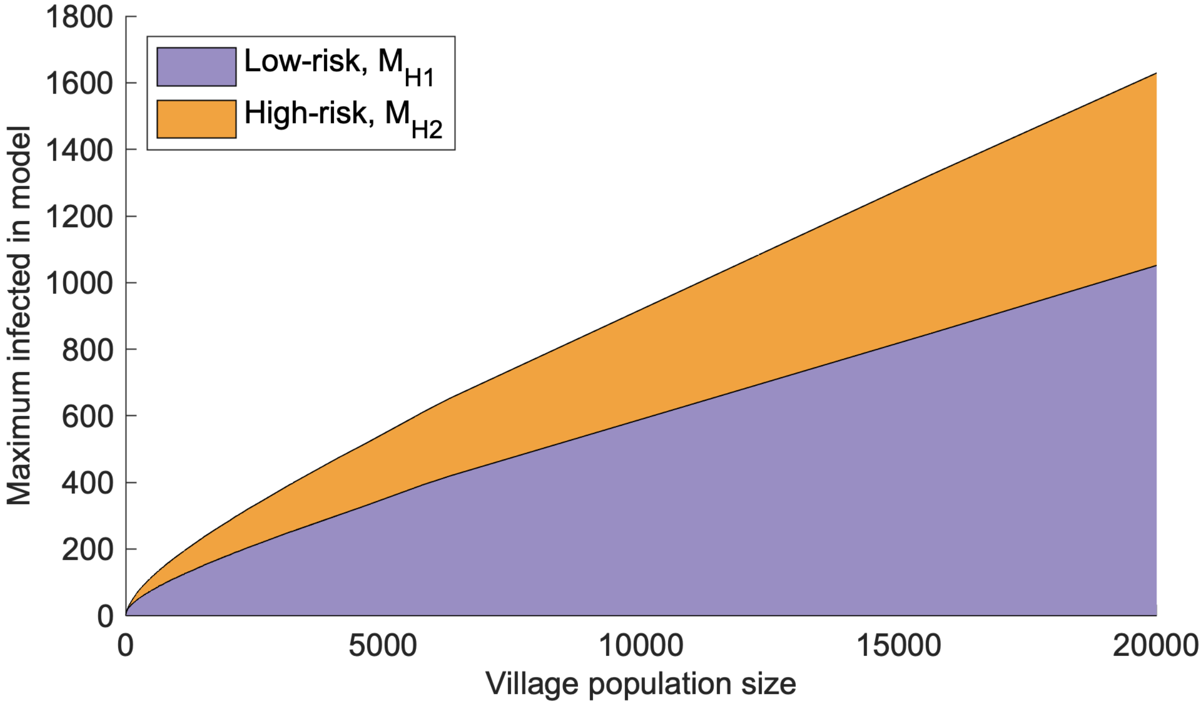


Figure S2: The imposed maxima of the number of infected low-risk people $M_L$, and high-risk people, $M_H$, to keep the state space sufficiently small.

Table S1: Parameter notation and values for suite of gHAT infection models. At the bottom are the parameters of the Kolmogorov forward equation model given in the first time point (in 1998) for the mean expected initial conditions of a village with population size $N_H = 1000$.

| Parameter | Description | Value | Source |
|---|---|---|---|
| $\mu_H$ | Natural human mortality rate | $5.4795 \times 10^{-5}$ days$^{-1}$ | [4] |
| $\omega_H$ | Human recovery rate | 0.006 days$^{-1}$ | [5] |
| $\sigma_H$ | Human incubation rate | 0.0833 days$^{-1}$ | [6] |
| $\phi_H$ | Stage 1 to 2 progression | 0.0019 days$^{-1}$ | [7, 8] |
| $\eta_H^{\text{post}}$ | Post-1998 exit rate from Stage 1 by treatment or death | 0.00012 days$^{-1}$ | [2] |
| $\eta_{H\text{amp}}$ | Maximum increase in the Stage 1 exit rate | 2.51 | [2] |
| $\gamma_H^{\text{pre}}$ | Pre-1998 exit rate from Stage 2 by treatment or death | 0.0017 days$^{-1}$ | [2] |
| $\gamma_H^{\text{post}}$ | Post-1998 exit rate from Stage 2 by treatment or death | 0.0019 days$^{-1}$ | [2] |
| $\gamma_{H\text{amp}}$ | Maximum increase in the Stage 2 exit rate | 0.514 | [2] |
| $d_{\text{steep}}$ | Steepness of the improvement rates for Stage 1 and Stage 2 passive detection | 0.941 | [2] |
| $d_{\text{change}}$ | Switching year of the improvement rate for Stage 1 and Stage 2 passive detection | 2006 | [2] |
| $N_H$ | Human population size | Varies | N/A |
| $\mu_V$ | Tsetse mortality rate | 0.03 days$^{-1}$ | [6] |
| $\sigma_V$ | Tsetse incubation rate | 0.034 days$^{-1}$ | [9, 10] |
| $\varepsilon$ | Reduced non-teneral susceptibility factor | 0.05 | [2] |
| $\alpha$ | Tsetse bite rate | 0.333 days$^{-1}$ | [11] |
| $m_{\text{eff}}$ | Effective tsetse density | 6.56 | [2] |
| $p_V$ | Probability of tsetse infection per single infective bite | 0.065 | [6] |
| $f_H$ | Proportion of blood-meals on humans | 0.09 | [12] |
| $K$ | Pupal density dependence | 111.09 | [13] |
| $\mathbb{P}$(pupating) | Probability of pupating | 0.75 | [13] |
| $\xi_V$ | Pupal death rate | 0.037 | [13] |
| $B_V$ | Total deposit rate | 0.0505 | [13] |
| Se | Sensitivity | 0.91 | [14] |
| Sp | Specificity | 0.9991 | [2] |
| $k_1$ | Proportion of low-risk humans | 0.902 | [2] |
| $k_2$ | Proportion of high-risk humans | 0.098 | [2] |
| $r$ | Relative bites taken on high-risk humans compared to low-risk | 6.61 | [2] |
| $u$ | Reporting probability for Stage 2 cases | 0.2737 | [2] |
| $\delta$ | Importation of infection rate in 2000 | $3.4 \times 10^{-6}$ days$^{-1}$ | [15] |
| $\zeta_1$ | Force of infection in low-risk group in Kolmogorov forward equation model | $1.303 \times 10^{-5}$ days$^{-1}$ | derived |
| $\zeta_2$ | Force of infection in high-risk group in Kolmogorov forward equation model | $8.525 \times 10^{-5}$ days$^{-1}$ | derived |
| $\psi_H$ | Recovery rate in Kolmogorov forward equation model | 0.001055 days$^{-1}$ | derived |

### Active screening

Active screening is where the population is tested for gHAT, typically with the Card Agglutination Test for Trypanosomiasis (CATT), or other rapid diagnostic test (RDT), with those people infected being treated (after disease confirmation and often stage determination). We simulate this process in the model by moving infected people directly from the infected class back to the susceptible class. This occurs through random selection from the low-risk population using a hypergeometric distribution $\mathrm{HG}(N_{H1}, c, k)$. We consider the number of infected people detected $c$ from the population $N_{H1}$ of low-risk people available to be screened, when a total of $k$ people are screened. Subsequently, we assume that there is a 91% probability of true positives being detected (the sensitivity of the screening algorithm). We use a binomial distribution $\mathrm{B}(c, \mathrm{Se})$, where the test sensitivity is $\mathrm{Se} = 91\%$ [14] to get the number of infected people screened that are identified.

We apply this process to the current probability vector for the population using the active screening matrix $\mathbf{A}$. This is a sparse lower-triangular transition matrix, which gives the new probability of being in each state after an active screening of given coverage. Thus $\mathbf{p}\,(Y, 0) = \mathbf{p}\,(Y-1, 365)\,\mathbf{A}\,(Y)$, i.e. the probability distribution at the start of the year is equal to the same distribution at the end of the previous year with the active screening matrix post-multiplied.

### Linking the villages

To consider the interactions of people moving between villages and becoming infected, causing importations of infection to their home village, as opposed to becoming infected through transmission in their village, we use the rate matrix $\mathbf{Q}_E$. We formulate this behaviour in two separate ways in the main manuscript. For

For the general set of villages in the health zone of Kwamouth, we assume a value of the rate of infectious importations from a previous study [16]. This rate decreases in time, where the rate is matched to the rate at which cases have decreased in the DRC [17].

However, when we consider the all of the actual villages of Kwamouth, we adapt the matrix $\mathbf{Q}_E$ by modifying this rate of infectious importations. We re-calculate the base rate of infectious importations at the steady state from 1998 to match the expected number of infected people to the expected number of infected people from the ODE formulation of the full model for the health zone with mixing in all villages. Then we remove the exponential decrease and replace this with a term proportional to the total number of expected infections in model predictions across all villages of a study region.

For a group of villages, we can calculate the total expected number of infected people by taking the sum of the expected number from each village $I_{\mathrm{Total}}$. In the rate matrix $\mathbf{Q}_E$, when considering an event that causes an importation of infection, we replace the term $\delta(N_{Hi} - I_{Hi})$, where $\delta$ is the rate of importations, with $\delta I_{\mathrm{Total}}(Y, y)/I_{\mathrm{Total}}(0)(N_{Hi} -$

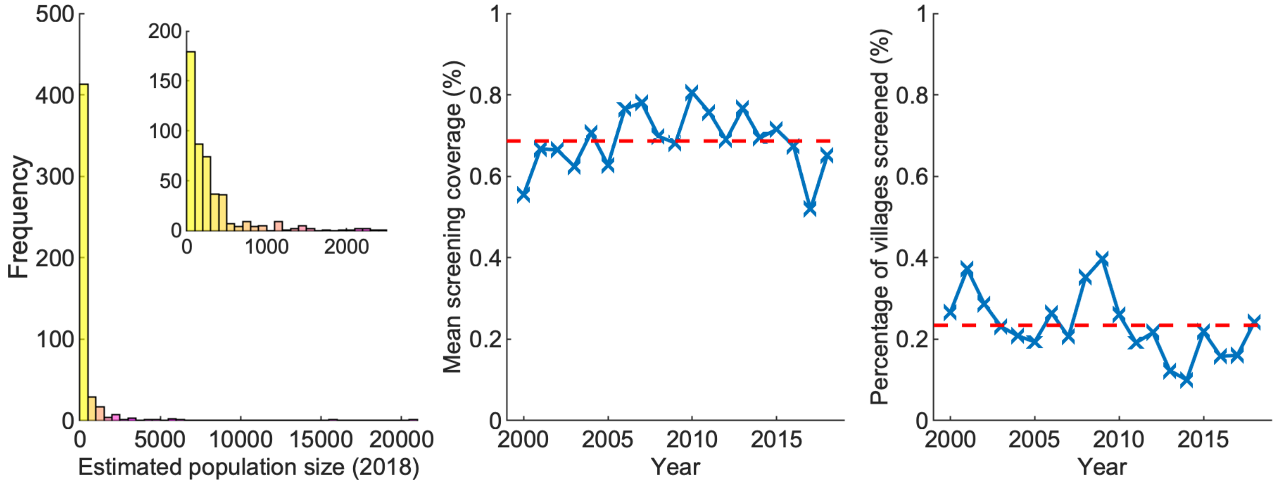


Figure S3: The villages of Kwamouth, DRC. (A) Histogram of the estimated population sizes of the villages from 2018, based on census data from the WHO HAT Atlas [3]. Inset plot shows smaller populations. (B) The annual mean screening coverage for a village when it is screened in Kwamouth (2000–2018). (C) The proportion of the villages of Kwamouth listed in the WHO HAT Atlas that are screened each year (2000–2018).

$I_{Hi}$). As such, for $Y = 1998$, we have the original rate, but as infection levels across all populations fall (or indeed increase), the importations of infection will fall (or increase).

In using this calculation we are making the simplification that $I_{\mathrm{Total}}(Y, y)$, the total expected number of infections across all the villages, is not dependent on the current state of a particular village, but formed simply from the total expected number across all villages. We do not want to compute the probabilities of all possible combinations of village states, as this would greatly increase the size of our system. This is assumption will be a good approximation for a large number of villages, although we note in making it we are neglecting second order effects. However, with this method, we have linked the infection dynamics in villages so that when there is no infection $I_{\mathrm{Total}}$ will be zero and so there will be no importations of infection and elimination of transmission will have occurred in the region.

### Population and active screening data

Population data for the health zone of Kwamouth is taken from the census data provided in the WHO HAT Atlas [3]. Here, where there are multiple census estimates for a village (all with the same geo-location), we calculate the time adjusted census estimate by assuming an annual population growth of 2.6% [17] and take the median value of the village population estimate as the population size of the village and then re-calculate the size in different years using the population growth rate. To keep the population size constant in the mode, we then assume the value from 2018. These calculations give a total population size for all villages in Kwamouth as 206,135, a value comparable with other sources [18].

Similarly, the village active screening data is taken from WHO HAT Atlas [3], where the number of people in each active screening event is reported. We derive the village-level active screening coverage by simply dividing the number of people screened by the population size of the village.

Active screening in Kwamouth has a mean screening coverage of 68.6% for each village-level active screening for the years 2000–2018 and 23.4% of years have an active screening for each village (Figure S3).

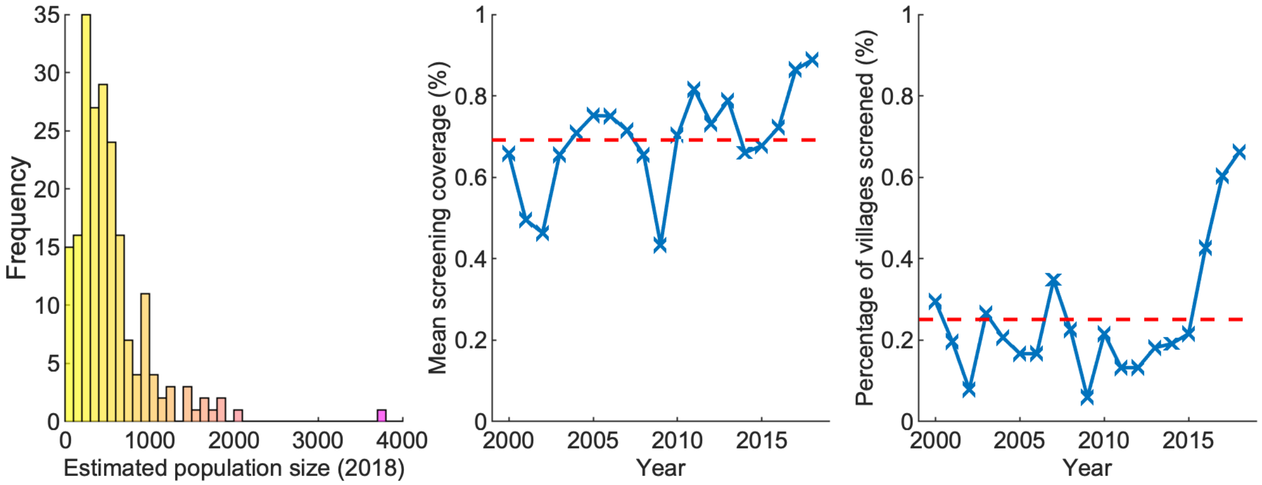


Figure S4: The villages of Mosango, DRC. (A) Histogram of the estimated population sizes of the villages from 2018, based on census data from the WHO HAT Atlas [3]. (B) The annual mean screening coverage for a village when it is screened in Mosango (2000–2018). (C) The proportion of the villages of Mosango listed in the WHO HAT Atlas that are screened each year (2000–2018).

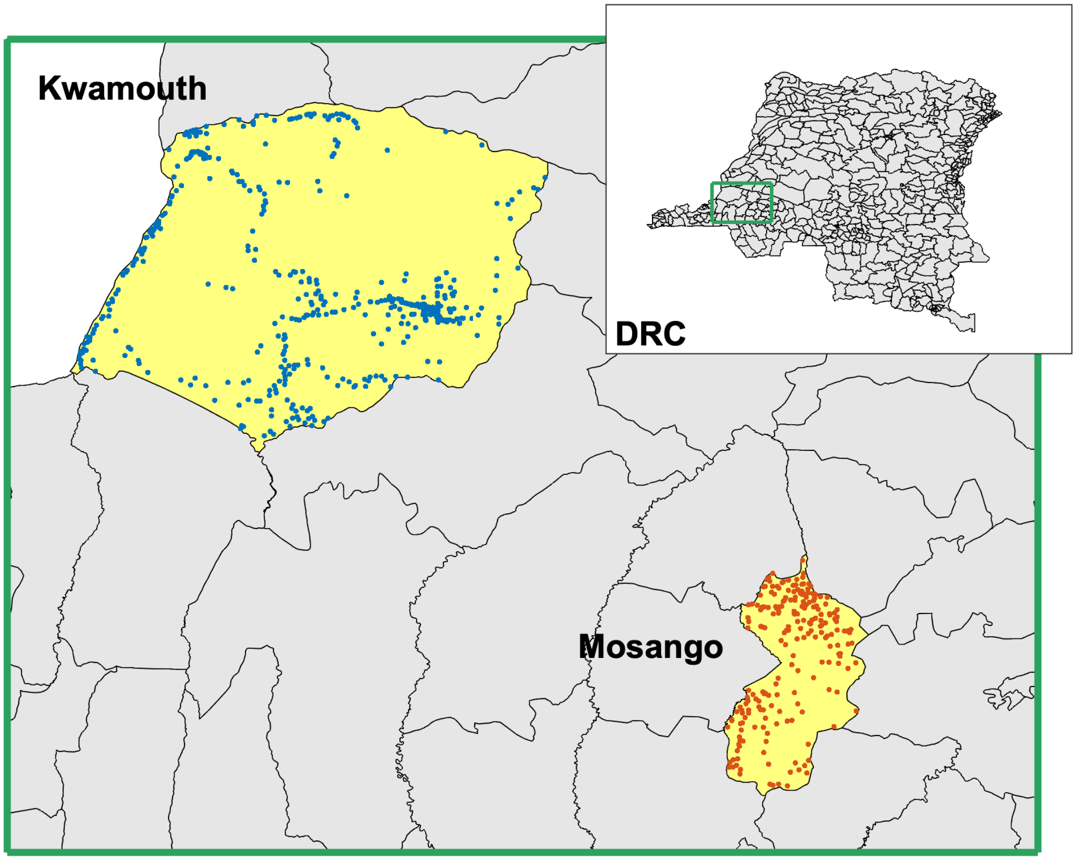


Figure S5: A map of the DRC. The health zones Kwamouth and Mosango are highlighted yellow and labelled. The villages of Kwamouth are shown by blue dots and the villages of Mosango shown by red dots. The inset map in the top right corner shows the location within the DRC.

#### Another health zone

The main manuscript primarily considers the dynamics of gHAT in a high-incidence health zone and this method could be applied to any health zone of the DRC. However, the specific parameterisation is derived from adapting the parameters (described in Equations 18 and 19) with values given by a fit to incidence data from the health zone of Kwamouth. We present an alternative parameterisation for the low-incidence health zone of Mosango (taken from Crump *et al.* [2] with similar screening behaviour (Figure S4) to produce similar results.

The locations of the health zones Kwamouth and Mosango within the DRC are shown in Figure S5. The villages of Kwamouth are shown by blue dots and the villages of Mosango shown by red dots. The active screening pattern in each health zone shows that larger villages tend to have more active screening events and fewer with very low coverage (Figure S6).

Figure S7 shows the infection distribution in the health zone of Mosango using the Kolmogorov forward equations in the same form as Figure 5 of the main manuscript. Results are qualitatively similar but without the large villages of Kwamouth. The maximum village population in Mosango in 3,721. There is higher probability of EOT in Mosango by 2030, at approximately 0.63 and 2029 is the expected year of EOT (shown in the main manuscript). These results are comparable to deterministic predictions [19].

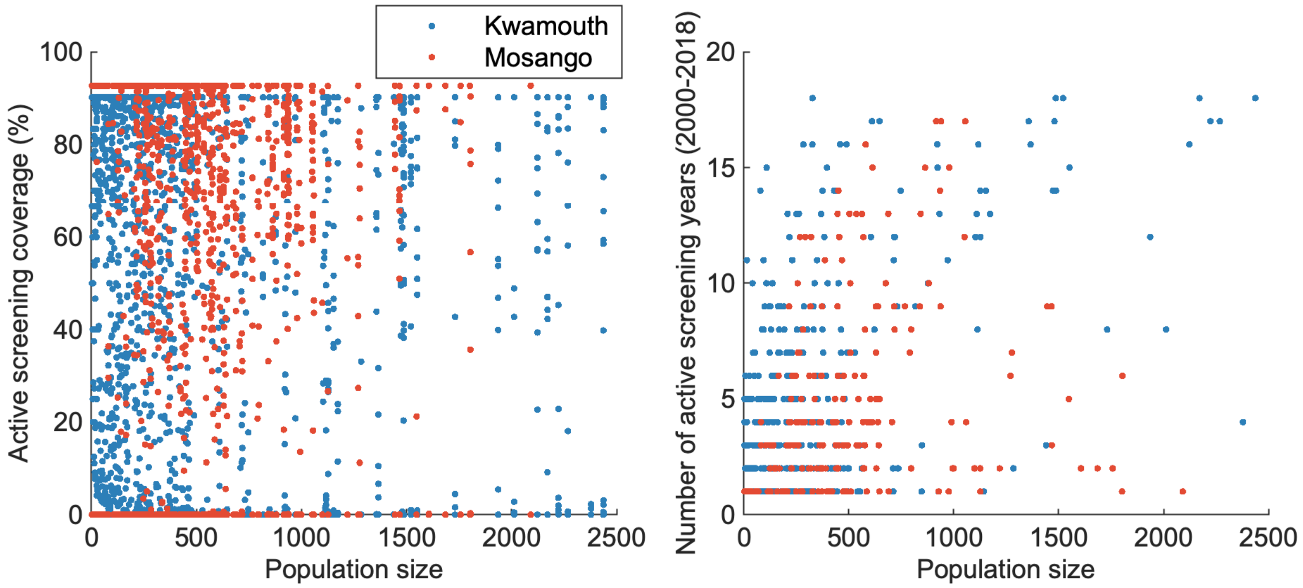


Figure S6: Active screening data by popuation size for Kwamouth and Mosango. The left plot shows the active screening coverag of each active screening event and the right plot show the number of times each village had an active screening in the years 2000–2018 (a maximum of 19).

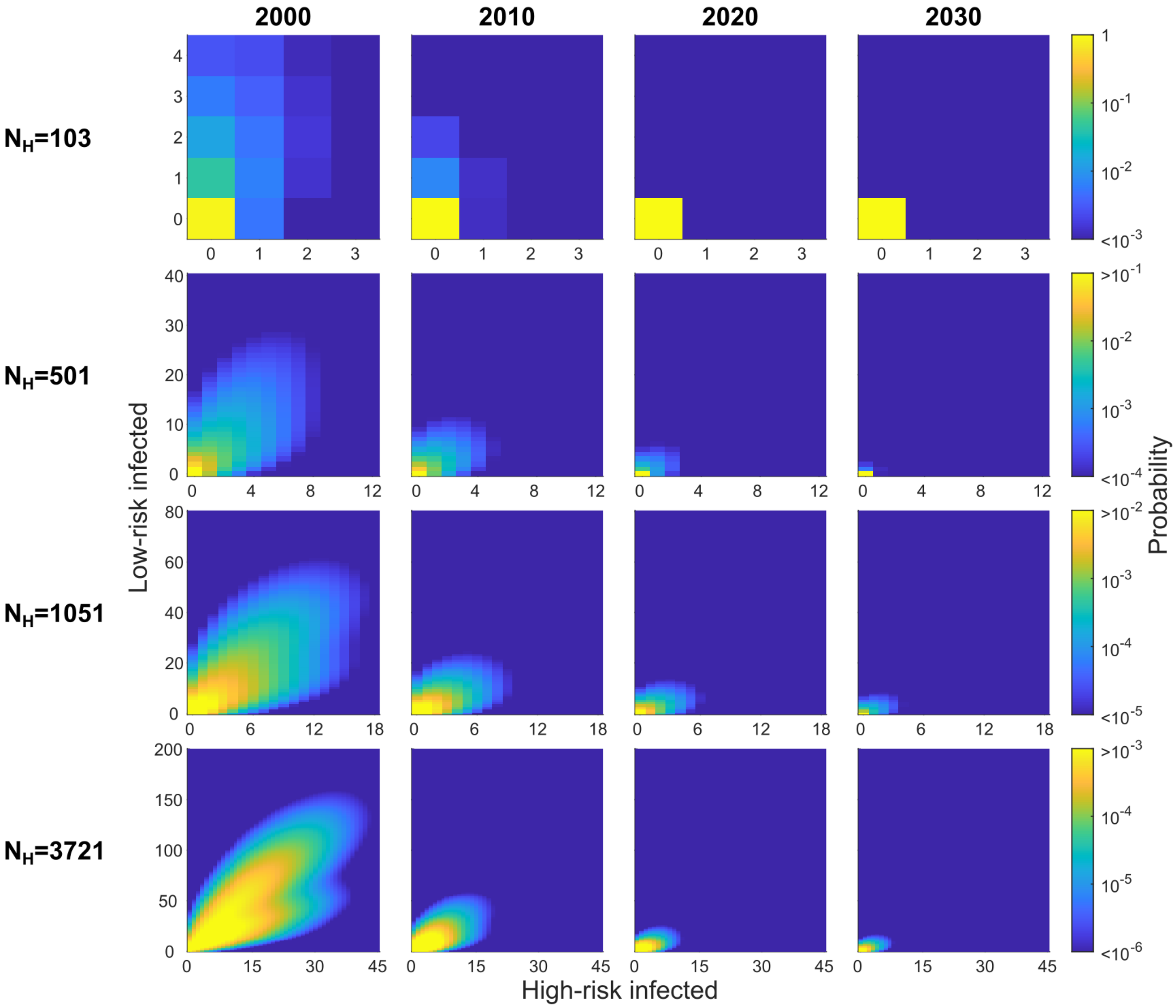


Figure S7: The risk distribution of infection in the villages of Mosango at selected time points. There are 204 villages with populations ranging between 3 and 3,721 of which we present the probability distribution of infected people in four villages ($N_H = 103$, $501$, $1,051$ and $3,721$).

#### Additional results

Due to lack of detailed data on when active screening occurs within a year, we have assumed that all active screening occurs on the first day of each day in all simulations. We now relax this assumption and consider 12 villages of population $N_H = 1,000$ of which one has an active screening at the beginning of each other (Figure S8). There is minimal difference between the infection dynamics under the two assumptions. There are marginally more infections when the screening is spread across the year, as there is benefit in reducing the infection numbers as much as possible at a given time to reduce transmission within the year. However, at the end of each year the infection levels are very similar, with bounce-back from the annual screening assumption. Thus, we can say that it is a reasonable approximation to assume all screening occurs on the first day of the year (or indeed any other day of the year). For modelling, assuming that all screening is done on the first day of the year is a useful simplification, since the available data will often not indicate exactly when active screenings occurred within a year and therefore it is useful to know that this lack of information will have minimal impact on predictions.

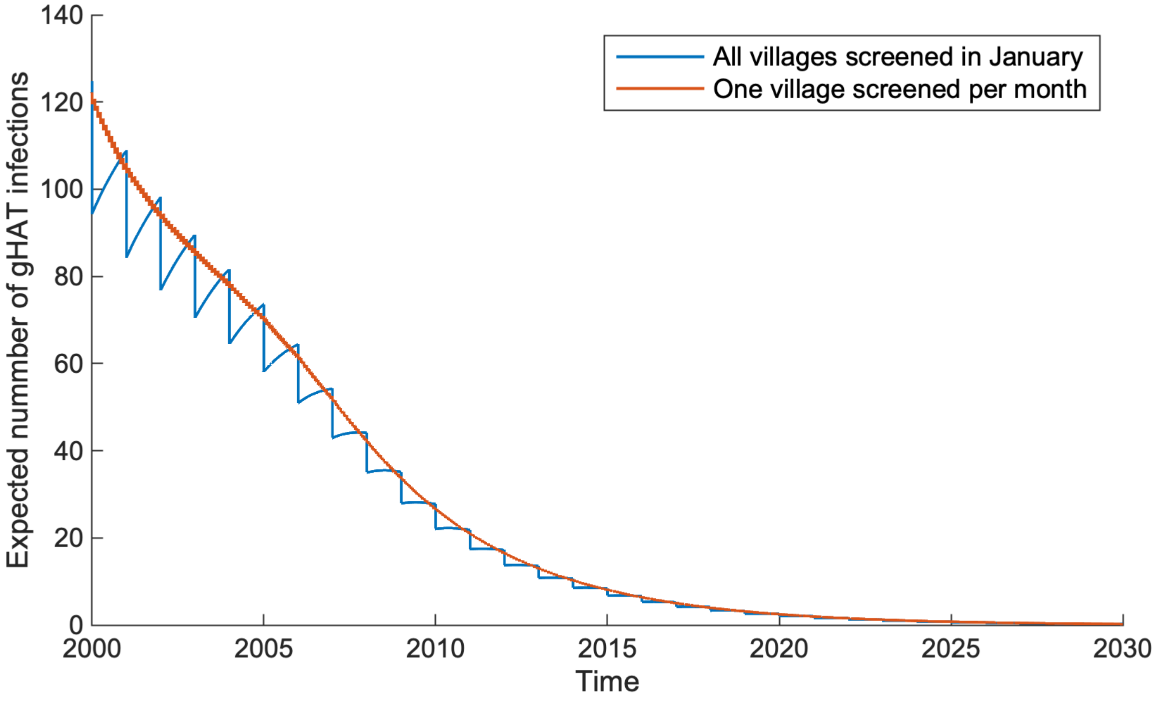


Figure S8: The expected number of gHAT infections for an area of 12 villages, each with a population of $N_H = 1,000$ people, given the assumption on timing of active screening: active screening in all villages occurs on the first day of the year, and one village is screened each month.

#### Comparison with ODE model

Comparing the results of the Kolmogorov forward equation model to the ODE variant we see very similar results, particularly for the Mosango health zone, which has a smaller population size. The prediction intervals also strongly overlap, giving confidence that these two model are showing very similar results. Hence, the Kolmogorov forward equation model, which significantly simpler, is a good approximation, but is fast to run compared to an event-driven stochastic approach, and provides the full and exact probability distribution of infection within the modelled health zone.

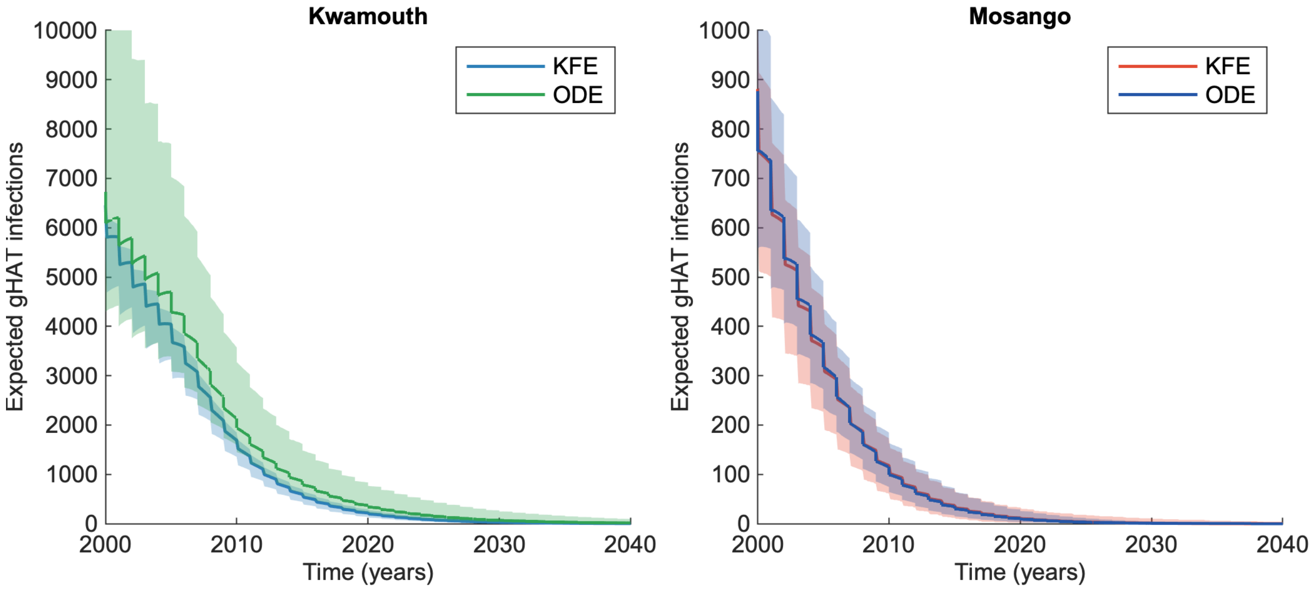


Figure S9: The expected number of infections in both Kwamouth and Mosango using the Kolmogorov forward equations (KFE) presented in the main manuscript and the model variant using ordinaring differential equations (ODE) from previous work [2, 19]. The both models shows 95% predictions intervals.

[4] The World Bank. Data: Democratic Republic of Congo. 2015.

[11] World Health Organization. Control and surveillance of human African trypanosomiasis. 2013.

[17] United Nations. World statistics pocketbook. 2016.

[18] DR Leasure, M Bondarenko, E Darin, and AJ Tatem. wopr: An R package to query the WorldPop Open Population Repository, version 0.4.0, 2020.

[19] Ching-I Huang, Ronald E Crump, Paul Brown, Simon EF Spencer, Erick Mwamba Miaka, Chansy Shampa, Matt J Keeling, and Kat S Rock. Shrinking the ghat map: identifying target regions for enhanced control of gambiense human african trypanosomiasis in the democratic republic of congo. *medRxiv*, 2020.
